# Nasopharyngeal Dysbiosis Precedes the Development of Lower Respiratory Tract Infections in Young Infants, a Longitudinal Infant Cohort Study

**DOI:** 10.1101/2021.10.13.21264939

**Authors:** Rotem Lapidot, Tyler Faits, Arshad Ismail, Mushal Allam, Zamantungwak T.H Khumalo, William MacLeod, Geoffrey Kwenda, Zacharia Mupila, Ruth Nakazwe, Daniel Segrè, W. Evan Johnson, Donald M Thea, Lawrence Mwananyanda, Christopher J Gill

## Abstract

**Background:** Infants suffering from lower respiratory tract infections (LRTIs) have distinct nasopharyngeal (NP) microbiome profiles that correlate with severity of disease. Whether these profiles precede the infection or a consequence of it, is unknown. In order to answer this question, longitudinal studies are needed.

**Methods:** We conducted an analysis of a longitudinal prospective cohort study of 1,981 Zambian mother-infant pairs who underwent NP sampling from 1-week through 14-weeks of age at 2-3-week intervals. Ten of the infants in the cohort developed LRTI and were matched 3:1 with healthy comparators. We completed 16S rRNA gene sequencing on the samples each of these infants contributed, as well as from baseline samples of the infants’ mothers, and characterized the normal maturation of the healthy infant NP microbiome, compared to infants who developed LRTI.

**Results:** The infant NP microbiome maturation was characterized by transitioning from *Staphylococcus* dominant to respiratory-genera dominant profiles during the first three months of life, similar to what is described in the literature. Interestingly, infants who developed LRTI had NP dysbiosis before infection, in most cases as early as the first week of life. Dysbiosis was characterized by the presence of *Novosphingobium, Delftia*, high relative abundance of *Anaerobacillus, Bacillus*, and low relative abundance of *Dolosigranulum*, compared to the healthy controls. Mothers of infants with LRTI also had low relative abundance of *Dolosigranulum* in their baseline samples compared to mothers of infants that did not develop an LRTI.

**Conclusions:** Our results suggest that NP microbiome dysbiosis precedes LRTI in young infants and may be present in their mothers as well. Early dysbiosis may play a role in the causal pathway leading to LRTI or could be a marker of other pathogenic forces that directly lead to LRTI.

**Funding:** This work was supported by The Southern Africa Mother Infant Pertussis Study – Nasopharyngeal Carriage (SAMIPS-NPC). PI Gill. Funder NIH/NIAID (1R01AI133080). WEJ and TF were supported by funds from the NIH, U01CA220413 and R01GM127430.

## Background

Lower respiratory tract infections (LRTI), including pneumonia and bronchiolitis, are the leading cause of death in children under five years of age, accounting for 1.3 million deaths each year, with 81% concentrated in children 2 years or younger (Cao et al., 2019; Fischer Walker et al., 2013). A necessary step leading to LRTI is the acquisition of a respiratory pathogen, such as *Streptococcus pneumoniae*. However, pneumococcal carriage is nearly universal among infants, only a few of whom develop severe invasive disease (Balsells et al., 2018; Yildirim et al., 2017, 2010). This indicates that the presence of the pathogen, while necessary, does not adequately address the more fundamental question of why some infants develop LRTI while most do not.

Increasingly, LRTI is seen as a consequence of the interaction between the pathogen and other contextual factors. Such factors include the net immune state of the host, intercurrent viral infections that may act transiently, or, in the case of HIV, for extended periods. Another factor may also be the microbial ecosystem in which the pathogen exists, *i*.*e*., the nasopharyngeal microbiome. Looking at the microbiome as an ecosystem model considers individual members of that ecosystem to exist in some dynamic equilibrium characterized by reciprocal loops of interaction. As such, the interaction between the microbiome and a specific potential pathogen (i.e., a pathobiont), could influence the behavior of that pathogen to either impede or promote LRTI (Brugger et al., 2016; Stewart et al., 2017).

In support of this ecosystem model, several cross-sectional studies have found that children with LRTIs often have distinct nasopharyngeal (NP) microbiome profiles at time of infection compared with healthy children. The NP microbiome profiles appear to be dominated by bacterial genera that differ between respiratory infections and health. For example, NP microbiomes dominated by *Streptococcus* and *Haemophilus* are associated with LRTI, whereas microbiomes profiles dominated by *Moraxella, Corynebacterium* and/or *Dolosigranulum* characterize healthy children. Further, NP microbiome characteristics correlate with the severity of respiratory disease and with clinical outcomes (de Steenhuijsen Piters et al., 2015; Hasegawa et al., 2017). While provocative, such observations largely rest on cross-sectional studies, and so cannot resolve the direction of cause and effect: we do not know whether these microbial profiles are a result of the infection or whether they preceded it. If the latter is true, then differences in the NP microbiome could potentially represent a state of vulnerability, participating in a causal pathway leading to LRTI.

To draw such inferences, it is necessary to have longitudinal data, with sampling of infants before the development of the LRTI. Since LRTI is a rare event, collecting longitudinal data is complicated by the large number of infants needed to be followed. Between 2015 and 2016, our team conducted a prospective cohort study and was able to create a biological sample library that allowed a longitudinal analysis of this kind. The study took place in Lusaka, Zambia, among 1,981 mother-infant pairs: The Southern Africa Mother Infant Pertussis Study – SAMIPS (Gill et al., 2016). The pairs were enrolled one-week post-partum. All enrolled infants were healthy and born term. At baseline, and every two-three weeks thereafter through 14 weeks of age, we obtained NP samples from mother and baby.

Within this cohort of 1,981 healthy infants a sub-set of 10 infants developed severe LRTI based on standard WHO clinical criteria (*Revised WHO classification and treatment of childhood pneumonia at health facilities • EVIDENCE SUMMARIES •*, n.d.). By comparing the infants who developed LRTI to matched healthy infants, we were able to conduct a time series analyses of NP microbiome of both infant populations, using 16S ribosomal DNA sequencing. We focused on the following fundamental analyses: 1) what is the ‘normal’ pattern of NP microbiome maturation over the first several months of life? 2) how does this contrast with the maturation of NP microbiome of infants who developed LRTI? 3) is there evidence that NP dysbiosis precedes the onset of LRTI? 4) are there distinct microbiome profiles that characterize sickness and health and other infant characteristics? 5) Is there also evidence of NP dysbiosis among the mothers of infants who later developed LRTI?

## Results

Within the SAMIPS cohort we identified ten infants who developed LRTI during the study period as defined by the WHO clinical criteria: cough, cold and fast breathing, chest indrawing or other general danger signs (lethargy, difficulty feeding, persistent vomiting, and convulsions) (*Revised WHO classification and treatment of childhood pneumonia at health facilities • EVIDENCE SUMMARIES •*, n.d.). We then matched these case infants by season of birth, number of siblings and HIV exposure status, with healthy comparators. With ten infants with LRTI and 3:1 matching, our analysis set consisted of 40 infants at ∼7 time points each. All infants were born healthy via vaginal delivery. Male sex was more common in infants who developed LRTI (p= 0.067). A third of infants with LRTI were born to mothers with HIV (receiving anti-retroviral treatment), compared to 40% of infants in the healthy group. Basic characteristics of the 40 infants are shown in **Table 1**. The symptoms and timing of sampling of the ten infants who developed LRTI are shown in **Table 2**.

**Table 1.** Characteristics of healthy infants and infants with LRTI

| Characteristics | Healthy Infants<br>(N=30) | Infants with LRTI<br>(N=10) | All Subjects<br>(N=40) | $p^*$ |
| --- | --- | --- | --- | --- |
| Sex, n (%) | | | | $p=0.067$ |
| Females | 16 (53.3%) | 2 (20.0%) | 18 (45.0%) |  |
| Males | 14 (46.7%) | 8 (80%) | 22 (55%) |  |
| Season of enrollment | | | | $p=0.224$ |
| Dry Season (May-Oct), n (%) | 28 (93.3%) | 8 (80.0%) | 36 (90.0%) |  |
| Rainy Season (Nov-Apr), n (%) | 2 (6.7%) | 2 (20.0%) | 4 (10.0%) |  |
| Median age at enrollment in days (IQR) | 7.0 (6 - 9) | 7.0 (6 - 10) | 7.0 (6 - 9) | $p=0.634$ |
| HIV exposed, n (%) | 13 (43.3%) | 3 (30.0%) | 16 (40.0%) | $p=0.456$ |
| Mean number of samples collected (SE) | 6.6 (0.2) | 6.6 (0.6) | 6.7 (0.2) | $p=0.958$ |

\*P values calculated using student t-test

**Table 2.** Clinical symptoms and age of 10 infants with LRTI at each study cisit/NP sampling

| Infants with LRTI | Sample Number (and age at sampling) |  |  |  |  |  |  |  |  |
| --- | --- | --- | --- | --- | --- | --- | --- | --- | --- |
|  | 1 | 2 | 3 | 4 | 5 | 6 | 7 | 8 | 9 |
| 1. | 7 days | 27 days | 42 days | 62 days | 79 days |  |  |  |  |
| 2. | 7 days | 27 days | 35 days | 42 days | 59 days | 73 days | 88 days | 104 days |  |
| 3. | 7 days | 11 days | 62 days |  |  |  |  |  |  |
| 4. | 7 days | 19 days | 45 days | 60 days | 68 days | 107 days |  |  |  |
| 5. | 7 days | 28 days | 42 days | 56 days | 69 days | 84 days | 100 days |  |  |
| 6. | 7 days | 21 days | 42 days | 56 days | 59 days | 73 days | 87 days | 96 days | 103 days |
| 7. | 7 days | 50 days | 59 days | 73 days | 87 days | 106 days |  |  |  |
| 8. | 7 days | 24 days | 27 days | 42 days | 61 days | 73 days | 90 days | 104 days |  |
| 9. | 7 days | 24 days | 39 days | 44 days | 65 days |  |  |  |  |
| 10. | 7 days | 23 days | 40 days | 61 days | 83 days | 99 days | 113 days |  |  |

- ☐ No symptoms ☐ Mild upper respiratory symptoms (cough/runny or blocked nose) ☐ LRTI symptoms (cough/runny or blocked nose with or without fever AND labored breathing/poor feeding/ inwarding of the chest/lethargy)

### 16S ribosomal DNA amplicon sequencing data and processing

We successfully sequenced 265 NP swabs from 40 infants, capturing a median of seven samples from each infant. The median age at first sampling was seven days, and the median age at final sampling was 104 days. We also sequenced two NP swabs from each infant’s mother at first and last time points, for a total of 345 samples from mothers and infants combined. In six of these samples, fewer than 10,000 reads aligned to RefSeq reference genomes and were excluded from further analysis. The remaining 339 samples had a median of 101,979 reads per sample assigned to reference genomes and were included in the analysis. From these, we detected 421 unique genera, spanning 14 unique phyla, which were assigned at least 100 sequence reads across all samples. Based on these results, we were confident in our ability to proceed with the ensuing analyses.

### Analysis One: What is the NP microbiome maturation in healthy infants in the first three months of life?

Given our ultimate goal of identifying characteristics of the NP microbiome in infants who develop LRTI, as a first step we describe the characteristics and evolution of NP microbiome of the healthy infants. We analyzed the NP samples from all of the infants who remained free of LRTI through the end of observation, using linear regression to track changes in relative abundance of genera over time spanning the period between enrollment after birth and 14 weeks of age.

We observed a stepwise pattern of maturation as the infants aged, summarized in **Figure 1a**, showing the relative abundance of different genera across each age averaged stratum. As can be seen, there is a clear shift in the abundance of dominant genera with time, with some dominating early in life, and others becoming more prominent as the children aged. Early in life, the dominant genera were *Staphylococcus* and *Corynebacteria*. According to a mixed-effects model, these genera declined in relative abundance as infants aged (*Staphylococcus*: p<10E-7, *Corynebacteria*: p<0.001) and were replaced primarily by *Streptococcus* (p<0.01 *Dolosigranulum* (p<0.001), *Moraxella* (p<0.001), and *Haemophilus* (p=0.02).

**Figure 1:**
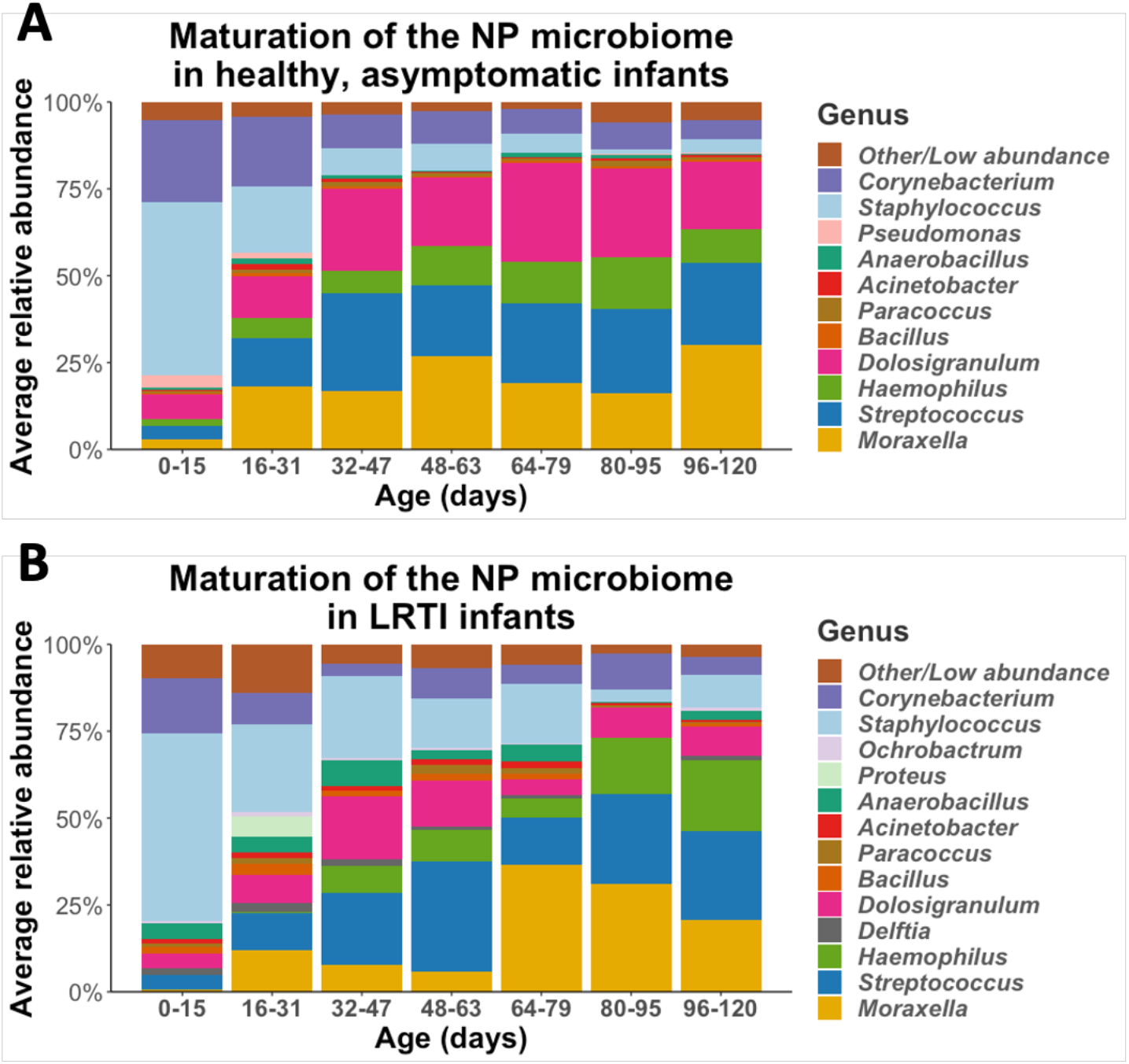
The maturation of the NP microbiomes of A) healthy, asymptomatic infants (n=30), and B) LRTI infants (n=10) over three months of observation. These stacked bar plots show the average relative abundance of the most common genera found in infant NPs, with samples binned by age.

We did not measure any significant change in the alpha diversity (richness within a given sample) of NP microbiomes as healthy infants aged, measured either by Shannon index (p=0.32) or Chao1 index (p=0.15). However, alpha diversity only reflects the number of dominant genera, and not whether the dominant genera are themselves diverse. Thus, when we clustered samples based on beta diversity (between sample diversity), measured as the Bray-Curtis dissimilarity between pairs of samples, we identified a distinct profile associated with samples from very young infants that contrasted against several profiles for more mature infant NPs (**Supplemental Figure 1a**). While each cluster is dominated by one or several of the most common genera, very few samples from healthy infants had high abundance of genera outside of the six most prominent genera. The primary axis of a Principal Coordinate Analysis (PCoA) (**Supplemental Figure 1b**) correlated with the age of the infants at the time of sampling, and stratified samples mainly by relative abundance of *Staphylococcus* and *Corynebacterium* in younger infants vs. the genera which were more common at older ages. The second PCoA axis distinguished between samples that were rich in *Moraxella* or *Dolosigranulum* from those rich in *Streptococcus* or *Haemophilus*. In summary, this analysis showed that the microbiomes of early infancy were highly dynamic over time, but that these shifts occurred in a structured and stereotypical pattern.

### Analysis Two: Does the maturation of the NP microbiome differ among infants who developed LRTI compared with healthy infants?

Given evidence from prior literature that during LRTIs the NP microbiome of infants is different than that compared to healthy infants, we set out to test whether the maturation of the NP microbiome in the first months of life is altered in infants who develop an LRTI. We repeated our analysis as described for healthy infants, stratifying into age groups and mapping the evolution of the NP microbiome over the first three months of life (**Figure 1b**). Infants who developed LRTI had similar general succession patterns as described for healthy infants, with high relative abundance of *Staphylococcus* early in life replaced by relative abundance of *Streptococcus, Haemophilus, Corynebacterium, Dolosigranulum* and *Moraxella*. Even though the general succession pattern of NP microbiome in infants with LRTI were similar to succession patterns of healthy infants, they exhibited distinct characteristics. Notably, the NP microbiome of infants who developed LRTI had, on average, higher relative abundance of specific genera including *Bacillus* (p=0.05) and *Delfia* (p<0.001) and lower relative abundance of *Dolosigranulum* (p<0.001).

As with the healthy control infants in our analysis 1, we did not observe any change in alpha diversity in LRTI infants as they aged (Shannon: p=0.08, Chao1: p=0.74). Analysis of the beta diversity between LRTI infant samples once again revealed a cluster of samples taken at very early time points, dominated by *Staphylococcus*, with samples taken from older timepoints exhibiting profiles rich in *Streptococcus, Dolosigranulum, Moraxella*, and *Haemophilus*. However, in LRTI infants we also observed a large sixth cluster, characterized by a high abundance of *Anaerobacillus* as well as various other rare genera (**Supplemental Figure 2**).

Nonmetric multidimensional scaling (NMDS) scaling of the beta diversity dissimilarity matrix between all samples allows us to visualize more holistic structural differences in the NP communities of healthy vs LRTI infants (**Figure 2**). When we project the age and eventual LRTI status of the infants into the NMDS ordination space, we can see that age correlates closely the primary NMDS axis, whereas LRTI status is mostly correlated with the secondary and tertiary axes, indicating differences in NP microbiomes between healthy and LRTI infants independent of the aging process.

**Figure 2:**
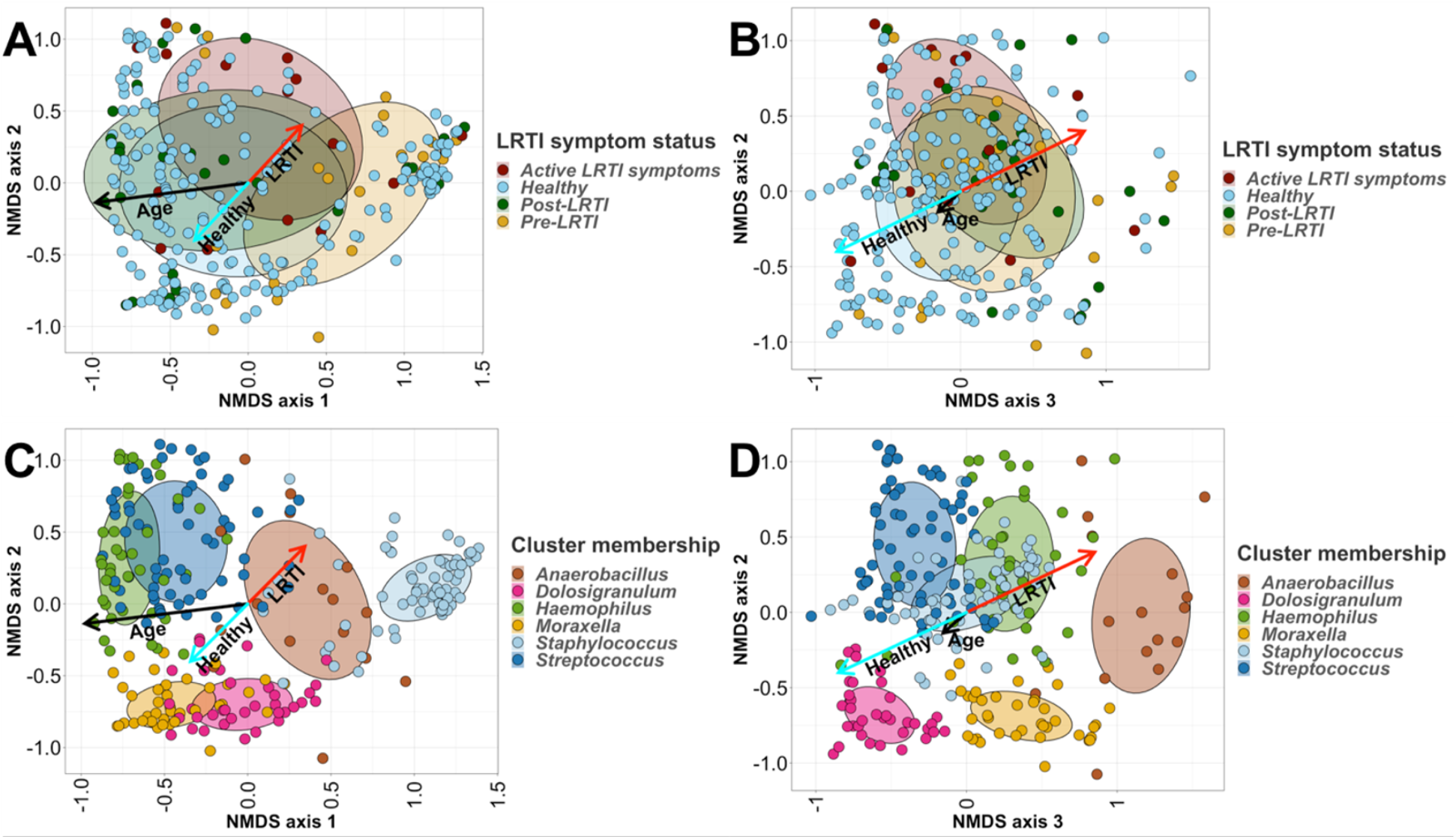
Nonmetric multidimensional scaling (NMDS) ordination plots of all infants’ (n=40) nasopharyngeal (NP) samples. We applied 3-dimensional NMDS ordination to the Bray-Curtis dissimilarity matrix between all infants’ NP swabs, and projected vectors into that ordination space representing the best fit correlations for the age at sampling (the black arrows) and LRTI status (the cyan arrows represent control infants, the red arrows represent LRTI infants). Plots A and C show the first two NMDS axes, while plots B and D show the second and third axes. Samples in plots A and B are colored based on their LRTI symptom progression, whereas samples in plots C and D are colored by their primary taxonomic profile cluster membership (see Figure 3 for details). Age is highly correlated with the first NMDS axis, and samples on the young end of the age vector mostly belong to the *Staphylococcus*-dominated profile, whereas samples on the older end tend to belong more to the *Haemophilus* and *Moraxella*-dominated profiles (A). The *Dolosigranulum*-dominated profile is associated with the healthy end of the vector for LRTI status, while the *Anaerbacillus*-dominated profile is associated with disease (A,C).

Since each infant developed an LRTI at a different age, stratifying the infants into age groups resulted in grouping together infants at different time points in relation to their disease – before the LRTI, at time of the LRTI, and following the LRTI. In order to describe maturation of the microbiome up to the time of LRTI, we created individual plots of the NP microbiome for each one of the ten sick infants (**Supplemental Figure 3**). These emphasize the high degree of heterogeneity of patterns over time across individuals.

### Analysis three: Is dysbiosis detectable at birth among infants who later develop LRTI?

To address this question, we performed analysis on the earliest NP samples taken from each infant at 7 days of age, comparing the microbiomes of those infants who eventually developed LRTIs to those who did not. At enrollment all infants were healthy by definition (based on enrollment inclusion/exclusion criteria), and therefore, infants who developed LRTI could collectively be grouped as “prior to infection” at that time point.

We used the R package DESeq2 to perform differential abundance tests. To be considered significant, a given genus would need to be differentially abundant with an FDR-adjusted p-value of less than 0.1 and also a mean relative abundance of at least 0.1% among either healthy or sick infants. We identified three options by which a genus could be different between the 2 groups: First, a genus that was identified exclusively in infants who developed LRTI, such as *Novosphingobium* (4/10). Second, genera that were more common in infants with LRTI (but were present in both groups), such as *Delftia* (8/10 in LRTI infants vs 13/30 in healthy infants). And third, there were genera that were detected in both groups, but were present with higher relative abundance in infants with LRTI compared to the healthy infants, such as *Anaerobacillus, Bacillus, Blastococcus, Brachybacterium, Ochrobactrum, Ornithinimicrobium*, and *Sphingomonas*. Overall, ten genera were significantly different in infants who later developed LRTI at the first time point (**Table 3**). Notably, *Dolosigranulum*, which has been identified in prior studies as being associated with a healthy microbiome, as was the case among the healthy infants here, had significantly lower relative abundance in infants who developed LRTIs than in healthy counterparts prior to the LRTI and even at the first sample time point.

**Table 3.** Differential abundance between control and LRTI infants at earliest observed timepoint

| Genus | Log Foldchange | Frequency in control infants | Frequency in LRTI infants | Adjusted p-value |
| --- | --- | --- | --- | --- |
| *Anaerobacillus* | 2.66 | 70% | 70% | 0.013 |
| *Bacillus* | 2.54 | 60% | 70% | <0.01 |
| *Blastococcus* | 5.36 | 0% | 10% | <0.01 |
| *Brachybacterium* | 5.22 | 3% | 30% | <0.01 |
| *Delftia* | 2.81 | 43% | 80% | <0.01 |
| *Dolosigranulum* | -4.14 | 57% | 50% | <0.01 |
| *Novosphingobium* | 6.80 | 0% | 40% | <0.01 |
| *Ochrobactrum* | 2.62 | 27% | 60% | <0.01 |
| *Ornithinimicrobium* | 4.77 | 3% | 20% | <0.01 |
| *Sphingomonas* | 2.72 | 17% | 40% | <0.01 |

### Analysis four: Are there distinct microbiome profiles that characterize sickness and health and other infant characteristics?

In order to identify specific microbial profiles, we applied hierarchical clustering to the Bray-Curtis dissimilarity matrix between each pair of samples from all infants. The Bray-Curtis dissimilarity matrix is a common tool in ecology for measuring the distance between different populations in terms of beta-diversity, and is bounded between 0 and 1, spanning ‘no dissimilarity’ to ‘complete dissimilarity’. We calculated at the genus level, using the *hclust* package in R. We used the Silhouette and Frey clustering indexes (NbClust) to determine the optimal heights at which to trim the dendrogram produced by the hclust function in R, splitting our samples into six primary clusters (Silhouette index) and 13 sub-clusters (Frey index). These six primary profiles were then named after the dominant genus within each cluster (the highest relative abundance genus). This yielded the following clusters: *Staphylococcus* dominant *Streptococcus* dominant, *Moraxella* dominant, *Dolosigranulum* dominant, *Haemophilus* dominant, and *Anaerobacillus* dominant profiles, corresponding to six of the seven most abundant genera across all our samples, as shown in **Table 4**. *Corynebacterium* is the only highly-abundant genus that does not compose the majority (or plurality) of relative abundance within any cluster; instead of being dominant in a subset of samples, *Corynebacterium* often co-occurred alongside the more dominant *Staphylococcus*, or to a lesser extent *Dolosigranulum*. For ease of reporting, we shall henceforth refer to each cluster by its most abundant genus.

**Table 4.** Relative abundance and frequency of the most common genera observed in the NP microbiome of healthy control and LRTI infants

| Healthy infants |  |  |  |
| --- | --- | --- | --- |
| Genus | Mean Relative Abundance | Frequency (Samples) | Frequency (Subjects) |
| *Streptococcus* | 19.4% | 93.4% | 100% |
| *Dolosigranulum* | 19.1% | 84.3% | 100% |
| *Moraxella* | 18.3% | 77.8% | 100% |
| *Staphylococcus* | 14.0% | 88.9% | 100% |
| *Corynebacterium* | 12.0% | 94.4% | 100% |
| *Paracoccus* | 0.95% | 66.7% | 100% |
| *Acinetobacter* | 0.84% | 66.2% | 100% |
| *Bacillus* | 0.55% | 52.5% | 100% |
| *Anaerobacillus* | 1.1% | 53.0% | 96.7% |
| *Pseudomonas* | 0.89% | 38.9% | 93.3% |
| *Delftia* | 0.32% | 37.9% | 90% |
| *Aeromonas* | 0.13% | 26.3% | 80% |
| *Haemophilus* | 8.7% | 38.4% | 73.3% |
| *Kocuria* | 0.11% | 18.7% | 63.3% |
| *Ochrobactrum* | 0.16% | 16.2% | 53.3% |
| *Escherichia* | 0.25% | 8.1% | 40% |
| *Enterobacter* | 0.18% | 9.6% | 40% |
| *Klebsiella* | 0.12% | 6.6% | 30% |
| LRTI infants |  |  |  |
| Genus | Mean Relative Abundance | Frequency (Samples) | Frequency (Subjects) |
| *Staphylococcus* | 22.1% | 90.8% | 100% |
| *Streptococcus* | 18.8% | 93.8% | 100% |
| *Moraxella* | 14.8% | 80% | 100% |
| *Dolosigranulum* | 9.6% | 58.5% | 100% |
| *Corynebacterium* | 8.3% | 86.1% | 100% |

Figure 3 shows the microbial composition of each of the 262 infant samples in our study which passed sample quality filters, grouped by the six primary profiles (**Figure 3A**) and the 13 sub-profiles (**Figure 3B**).

**Figure 3:**
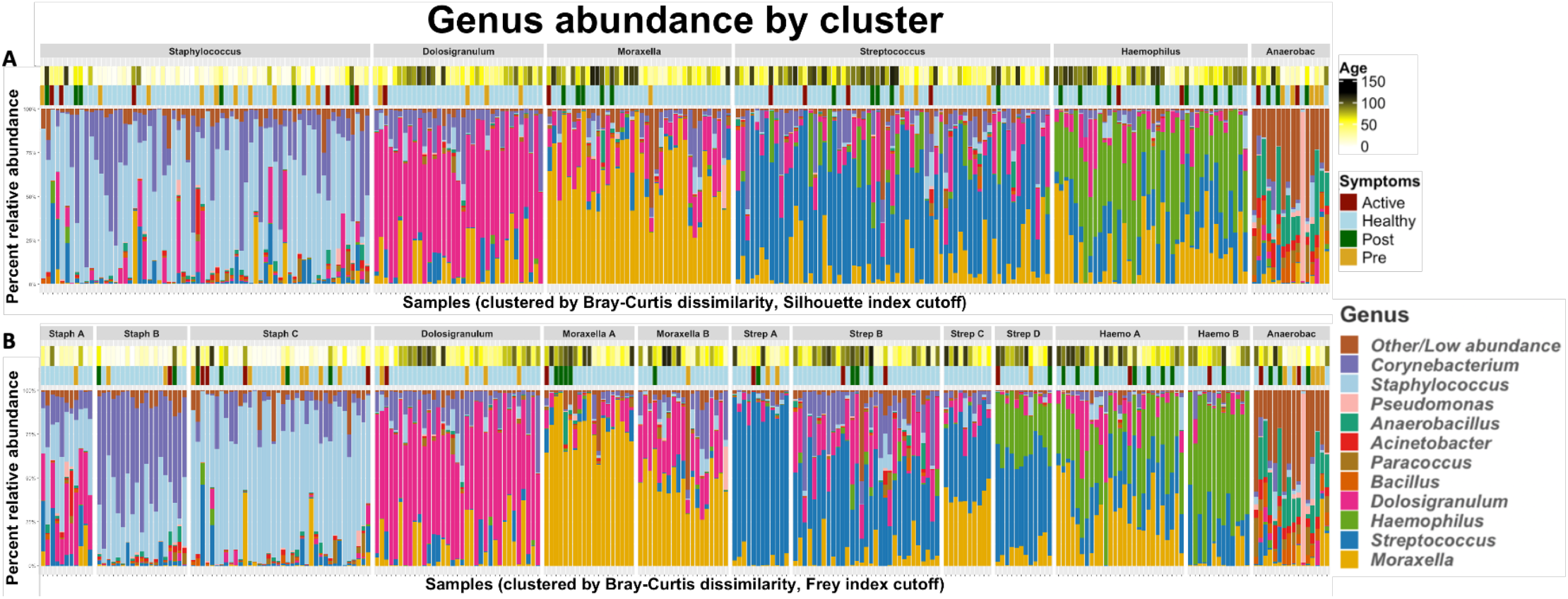
The taxonomic profiles of all infant NP samples (n=40), clustered by pairwise Bray-Curtis dissimilarity. Clusters were defined by performing hierarchical clustering on the beta diversity matrix and then cutting the resulting dendrogram into an optimal number of clusters according to the A) Silhouette index (6 clusters) and B) Frey index (13 clusters). The color bars above the stacked bar plots indicate the infants’ ages at the time of each sample and their LRTI status – “healthy” indicates an infant which did not develop LRTI symptoms during our observation.

Fisher’s exact tests revealed that the *Anaerobacillus* dominant profile was highly associated with infants who developed LRTIs, (p<0.01, estimated odds-ratio=5.74). The *Staphylococcus* sub-profile Staph-C was associated with LRTI infants (p=0.04, estimated odds-ratio=2.26), and the *Streptococcus* subcluster Strep-C (which is also rich in *Moraxella*) was associated with healthy infants (p=.0.07). Using ANOVA to assess the association of each profile with age, the *Staphylococcus* dominant profile was clearly associated with samples from younger infants compared to all other profiles, and the *Anaerobacillus* dominant profile was associated with younger samples when compared to the *Haemophilus* and *Streptococcus* profiles (**Table 5**).

**Table 5.** Associations between NP microbiome profiles with LRTI status and age

| Associations with LRTI status |  |  |  |
| --- | --- | --- | --- |
| Cluster/Subcluster | Odds ratio estimate | Odds ratio range | P-value |
| Anaerobacillus | 5.74 | 1.80-20.11 | <0.01 |
| Staphylococcus C | 2.26 | 1.02-4.92 | 0.04 |
| Streptococcus C | 0.00 | 0.00-1.34 | 0.07 |
| Associations with age (in days) |  |  |  |
| Cluster | Cluster | Difference in age | Adjusted P-value |
| Staphylococcus | Moraxella | 39 | <0.01 |
| Staphylococcus | Dolosigranulum | 52 | <0.01 |
| Staphylococcus | Streptococcus | 41 | <0.01 |
| Staphylococcus | Haemophilus | 44 | <0.01 |
| Staphylococcus | Anaerobacillus | 21 | 0.05 |
| Anaerobacillus | Streptococcus | 20 | 0.1 |
| Anaerobacillus | Haemophilus | 24 | 0.05 |

We visualized the association between LRTI status and NP taxonomic profiles using NMDS ordination (**Figures 2C and 2D**). By projecting infants’ LRTI status and age into the ordination space, we can see that the vector corresponding to healthy samples points towards the *Dolosigranulum* profile (and to a lesser extent towards the *Moraxella* profile), while the LRTI vector points towards the *Anaerobacillus* profile.

Together, these results reinforce a number of our previous observations; in particular, we can see that there is a general trend for infant NP microbiome profiles to shift from being *Staphylococcus* dominant shortly after birth towards several other profiles. We also see a clear dysbiotic pattern, comprising higher than normal relative abundance of *Anaerobacillus* as well as higher prevalence of rare genera which typically make up an extremely low portion of (or are completely absent from) healthy NP microbiomes.

### Analysis five: Is the NP microbiome of mothers of infants who develop LRTI different than mothers of healthy infants?

Observing distinct characteristics of the NP microbiome of infants as early as age 7 days, suggested that these profiles might be related to in-utero exposures, transmittable immunologic factors, and/or host genetics. That led us to question whether mothers of infants who develop LRTI have themselves distinct characteristics of the NP microbiome. We analyzed the first NP swabs from each of the mothers enrolled in our study taken at the infants’ day seven enrollment visits, correlated their microbiomes to those of their infants, and used DESeq2 to establish which genera were differentially abundant between mothers of LRTI infants and mothers of healthy infants. Similar to the pattern seen in the infants themselves, the mothers of infants who developed LRTIs had significantly decreased relative abundance of *Dolosigranulum* (p=0.05) as compared to mothers of healthy infants at 7 days of infant’s life.

## Discussion

In this analysis, we show that the NP microbiome of infants with LRTI differs from that of healthy infants and that there is clear evidence of dysbiosis preceding the onset of LRTI. Intriguingly, we observed different microbiome patterns in the mothers of infants who later developed LRTI and those whose children remained healthy. That, and the fact that the microbiome of mother-infant pairs is more closely correlated within pairs than across pairs, suggests that some of the infant dysbiosis has transgenerational origins. As an overall synthesis, our data suggest that there are quantitative and qualitative differences between infants (and their mothers) who do and do not develop LRTI. This supports the hypothesis that LRTI is not a random event, but rather may reflect predispositions that are generally unobserved but may nonetheless play an essential or contributory role in the pathogenesis of childhood LRTI.

The nasopharynx is the ecologic niche of respiratory pathobionts, and in this ecosystem they will either become invasive or remain merely colonizers. The NP microbiome at time of infection is associated with the risk of development of LRTI and its severity. But there is also good reason to believe that the maturation of the NP microbiome in the first months of life, and not only its characteristics at time of infection, is associated with respiratory health and development of disease later in life. For example, maturation of the gut microbiome is known to regulate the immune system evolution and is associated with the development of diseases later in life such as obesity and type 1 diabetes (Bokulich et al., 2016; Stewart et al., 2018). Gut microbial dysbiosis in children often predisposes to recurrent *C. difficile* infections (Ihekweazu and Versalovic, 2018). Thus, a similar association between the NP microbiome and risk of respiratory infections is a plausible theory for which there is precedent.

We have shown that we can characterize the normal, healthy maturation of the NP microbiome over the first months of life, and how this maturation is different in infants who develop early LRTI. While the evolution of the normal microbiome is highly dynamic, it proceeds in a stereotypical fashion, with stepwise shifts from a flora dominated by skin organisms (*Staphylococcus)*, to one that is dominated by genera more typically associated with the respiratory tract (*Dolosigranulum, Streptococcus, Haemophilus and Moraxella)*. Similar microbial succession patterns were previously described in other birth cohorts (Biesbroek et al., 2014).

By contrast, infants who develop LRTI have similar general succession patterns as healthy infants; transitioning form high relative abundance of *Staphylococcus* to high relative abundance of genera associated with the respiratory tract, but unlike healthy infants, the evolution of their NP microbiome is characterized by low relative abundance of specific genera associated with ‘health’, such as *Dolosigranulum*, and high relative abundance of other genera that appear unique, such as *Anaerobacillus, Bacillus*, and a mixture of ‘other’ uncommon genera. Additionally, the LRTI infants’ microbiomes include a larger number of uncommon and transient genera, presenting a picture that is more chaotic than what is seen in the healthy infants.

Case-control studies have consistently demonstrated an association between NP microbiome characteristics and LRTIs at time of disease, though interpretation in terms of causality could not be shown. The relatively high abundance of *Dolosigranulum/Corynebacterium* and *Moraxella* are correlated with healthy states (Mansbach et al., 2016), whereas NP microbiomes enriched with *Streptococcus* and *Haemophilus* are associated with LRTI’s that also correlate with severity of disease (de Steenhuijsen Piters et al., 2016; Kelly et al., 2017). But are these microbial profiles a result of the infection? Or were they present before the infection?

We were able to identify several microbiome profiles which appear to cluster by chronological age, LRTI and health. Our results indicate that young infants who developed LRTI, had NP microbiome dysbiosis prior to acquiring the infection, and as early as 7 days of life. These infants have NP microbiome enriched with *Aneorobaccillus/Bacillus, Acinetobacter*, and other uncommon/unspecified genera, and also have relatively lower abundance of *Dolosigranulum*. Our intriguing results suggest that their mothers NP microbiome at the same early time point also differed from that of mothers of healthy infants.

The interaction between host, microbiome and pathobionts is complex and most probably multidirectional. The NP microbiome, known to be associated with environmental factors (breastfeeding, mode of delivery) (Bosch et al., 2017; Brugger et al., 2016). could also very well be a reflection or marker, of host genetics and immune system function, which would explain why so early in life “high risk” profiles are observed. New acquisition of a pathobiont in the nasopharynx initiates interactions between the pathobiont and other organisms residing in the nasopharynx. These interactions modify metabolic activity and gene expressions of the pathobiont that influence whether the pathobiont becomes invasive. The interactions themselves between organisms in the nasopharynx also modify host immune response which underscores the complex relationship between host, microbiome and pathogens (de Steenhuijsen Piters et al., 2019).

The key unresolved question is what role dysbiosis plays in the causal pathway leading to pneumonia: is dysbiosis a marker of other unobserved forces that lead to pneumonia, such as underlying host genetic or immunologic factors? Or does dysbiosis play a role in the causal pathway leading to LRTI? While our data cannot resolve this question, the implication of our findings are substantial. Our findings suggest that NP dysbiosis identified in the first days of life is associated with higher risk of developing LRTI in early infancy. This suggests that there is an important window of opportunity for identifying these infants and intervene. According to our findings, it may even be that we can identify these infants, by examining the mothers.

Our study has several limitations. Infants were followed until the age of three months, and thus our findings could not be generalized to older age groups. On the other hand, it is possible that infants included in our healthy control group developed LRTI after the study period, in that case our results are biased towards the null, possibly underestimating differences between the two groups.

A further limitation is that we do not know the causative pathogen of the LRTIs, and whether these were viral, bacterial, or mixed pathogen LRTIs. LRTI is a heterogeneous set of conditions, and it is plausible that dysbiosis can interact in pathogen-specific ways. The diagnosis of LRTI was based only on clinical data. Even though different pathogens interact in different ways with the NP microbiome and the host immune system, our data suggests that there is a common NP microbiome risk profile, regardless of the causative pathogen. Lastly, while our analysis included a very large number of longitudinal samples, our sample size only included 10 infants who developed LRTI. However, LRTI is a comparatively rare event and requires longitudinal surveillance of thousands of subjects over an extended period to identify even a few cases, which accounts for the paucity of research on this topic. Logistically, it is immensely challenging and resource intense to create and sample a cohort in the way we have done. Nonetheless, further research will be needed to confirm or refine these initial observations. If confirmed, these findings are not only critical to our understanding of factors that lead to the development of LRTI, and why one infant develops an LRTI while others do not, it also suggests that we have a window of opportunity to identify these “at-risk” infants before their infection, and to potentially intervene. These prevention measures could have a high impact on decreasing burden of LRTI in infancy.

## Conclusions

Dysbiosis of the NP microbiome in infants precedes LRTIs, suggesting at minimum a signal of infants at higher risk for LRTIs, and possibly a causative role in the development of these infections. Specific NP microbiome profiles which could be identified perinatally, and appear to be associated with a higher risk of developing LRTIs in early infancy, present a potential window of opportunity for interventions. Our findings should be confirmed by large scale longitudinal studies.

## Materials and Methods

### Study population

This is a nested time-series case comparator study within the prospective longitudinal Southern Africa Mother-Infant Pertussis study (SAMIPS). SAMIPS was a study conducted in Zambia in which infants and their mothers were followed over the first 3 months of life. Full methods description is previously detailed by Gill et al (Gill et al., 2016), in short: All infants enrolled to SAMIPS were less than ten days of age, born term, via normal vaginal delivery, and deemed healthy after birth. All infants received scheduled vaccines. Written informed consent was obtained as appropriate from mothers of infants enrolled in the study.

The study was approved by the ethical review committees at the ERES Converge IRB in Lusaka, Zambia, and at Boston University Medical Center. All mothers provided written informed consent, with consent provided in English, Bemba or Nyanja as preferred by the participant.

### Study design

Mother-infant pairs were enrolled when mothers returned for their first postpartum well-child visit at one week of age. At enrollment, and 2-3-week intervals thereafter, through 14 weeks, we obtained a posterior nasopharyngeal (NP) swab from both mother and baby, with additional swabs obtained adventitiously if either returned seeking care for an acute respiratory infection.

Within the SAMIPS cohort, we identified ten infants who during the study period suffered from symptoms of lower respiratory tract infection (LRTI) as adopted from the WHO (*Revised WHO classification and treatment of childhood pneumonia at health facilities • EVIDENCE SUMMARIES •*, n.d.). Sick infants were matched 1:3 with healthy comparators by season of enrollment, maternal age and household composition.

### Sample processing and storage

NP swabs were obtained from the posterior nasopharynx using a sterile flocked tipped nylon swab (Copan Diagnostics, Merrieta, California). The swabs were then placed in universal transport media, put on ice and transferred to our onsite lab on the same campus, where they were aliquoted and stored at -80°C until DNA extraction. DNA was extracted using the NucliSENS EasyMagG System (bioMérieux, Marcy l’Etoile, France). Extracted DNA was stored at our lab located at the University Teaching Hospital in Lusaka at -80°C. Sample collection, processing and storage were previously described (Gill et al., 2016).

### 16S ribosomal DNA amplification and MiSeq sequencing

For 16S library preparations, two PCR reactions were completed on the template DNA. Initially the DNA was amplified with primers specific to the V3–V4 region of the 16S rRNA gene (Klindworth et al., 2013). The 16S primer pairs incorporated the Illumina overhang adaptor (16S Forward primer 5’-TCGTCGGCAGCGTCAGATGTGTATAAGAGACAGCCTACGGGNGGCWGCAG-3’; 16S reverse primer 5’-GTCTCGTGGGCTCGGAGATGTGTATAAGAGACAGGACTACHVGGGTATCTAATCC-3’) Each PCR reaction contained DNA template (∼12 ng), 5µℓ forward primer (1μM), 5 µℓ reverse primer (1μM), 12.5 µℓ 2 X Kapa HiFi Hotstart ready mix (KAPA Biosystems Woburn, MA), and PCR grade water to a final volume of 25µℓ. PCR amplification was carried out as follows: heated lid 110°C, 95°C for 3 min, 25 cycles of 95°C for 30s, 55°C for 30s, 72°C for 30s, then 72°C for 5 min and held at 4°C. Negative control reactions without any template DNA were carried out simultaneously.

PCR products were visualized using Agilent TapeStation (Agilent Technologies, Germany). Successful PCR products were cleaned using AMPure XP magnetic bead-based purification (Beckman Coulter, IN). The IDT for Illumina Nextera DNA UD Indexes kit (Illumina, San Diego, CA) with unique dual index adapters were used to allow for multiplexing. Each PCR reaction contained purified DNA (5 μℓ), 10 μℓ index primer mix, 25 μℓ 2X Kapa HiFi Hot Start Ready mix and 10 μℓ PCR grade water. PCR reactions were performed on a Bio-Rad C1000 Thermal Cycler (Bio-Rad, Hercules, CA) Cycling conditions consisted of one cycle of 95°C for 3 min, followed by eight cycles of 95°C for 30 s, 55°C for 30 s and 72°C for 30 s, followed by a final extension cycle of 72°C for 5 min.

Prior to library pooling, the indexed libraries were purified with Ampure XP beads and quantified using the Qubit dsDNA HS Assay Kit (Thermo Fisher Scientific, Waltham, MA). Purified amplicons were run on the Agilent TapeStation (Agilent Technologies, Germany) for quality analysis before sequencing. The sample pool (2 nM) was denatured with 0.2N NaOH, then diluted to 4 pM and combined with 10% (v/v) denatured 20 pM PhiX, prepared following Illumina guidelines. Samples were sequenced on the MiSeq sequencing platform at the NICD Sequencing Core Facility, using a 2 × 300 cycle V3 kit, following standard Illumina sequencing protocols.

Sequencing data were processed using QIIME2 (Bolyen et al., 2019) and Pathoscope2 (Hong et al., 2014). Samples with less than 10,000 reads were excluded from further analysis.

### Data processing

We assessed the quality of the sequencing data using FastQC (Andrews, 2010), which indicated that the overall sequencing quality was excellent, with mean Phred quality scores remaining greater than 30 (>99.9% accuracy) for over 200bp for both forward and reverse reads. We used *Trimmomatic* (Bolger et al., 2014) to trim Illumina adapters and remove low-quality sequences, setting the tool’s parameters to LEADING:6, TRAILING:6, SLIDINGWINDOW:6:15, and MINLEN:36. This quality filtering removed less than 0.5% of reads from each sample.

We used PathoScope 2 to assign sequencing reads to bacterial genomes. We used all of RefSeq’s representative bacterial genomes (downloaded November 2, 2018) as a PathoScope reference library. From PathoScope’s subspecies-level final best hit read numbers, we compiled counts tables and relative abundance tables for each sample at the phylum, genus, and where possible, to the species level.

### Data and statistical analysis

#### NP microbiome characteristics and evolution over time

We describe the normal evolution of the NP microbiome in healthy infants over the first three months of life. We calculated microbial richness using Chao1 index, and diversity of microbial taxa using the Shannon diversity index. We report the individual evolution of NP microbiome of each of the 10 infants who develop LRTI. In order to establish statistical significance, we used the *lmer* function from the *lme4* package for R (Bates et al., 2015) to apply a mixed-effects linear model to the log counts per million (logCPM) value of each genus, including age and HIV exposure as fixed effects and the study subject as a random effect. All p-values generated by these linear models are reported after False Discovery Rate (FDR) adjustment for multiple comparisons using the Benjamini-Hochberg method (Benjamini and Hochberg, 1995). We only generated mixed-effects models for genera which had an average relative abundance of at least 0.5% across all healthy infant samples.

For visualization of the development of healthy NP microbiota, we grouped all infant samples by age (in days) into 7 bins, each comprising a 16-day age window (0-15 days, 16-31 days, etc). We only visualized genera which had an average relative abundance of at least 1% across all samples. The relative abundances of all genera which did not meet this threshold were summed into a group labelled “Other/Low abundance” for plotting purposes only.

We calculated estimates of the alpha diversity within each sample based on the species-level counts tables generated by PathoScope 2. We calculated alpha diversity using two methods: the Chao1 index, which estimates the total number of species present within a sample, and the Shannon index, an entropy-based metric which incorporates both the number of species present and the evenness of abundance among those species. The Chao1 index was calculated using the R package *fossil* (Vavrek, 2011) and the Shannon index was calculated using the R package *vegan* (available via CRAN) (Oksanen et al., 2019) each with a rarefaction depth of 10,000. We constructed a mixed-effects linear model as described above, except using each alpha diversity metric as a response variable, in order to test whether alpha diversity changed as infants aged.

#### Analysis of the association between the NP microbiome and the development of LRTI

We used the lmer function from the *lme4* package (described above) to build mixed-effects linear models to compare the development of the NP microbiomes of infants who developed LRTIs to those of healthy infants. This time, we included infection status and the interaction of infection status with age as fixed-effect covariates in addition to age and HIV exposure, as well as study subject as a random effect. Once again, p-values were generated using the Anova function of the *car* package (Weisberg, 2019) and then FDR corrected.

We similarly modified the models we had used to test alpha diversity in order to see if either Shannon or the Chao1 index values were different in LRTI infants, once again adding infection status and the interaction between infection status and age as fixed effects.

#### Differential abundance analysis at first timepoints

We performed differential abundance between the first samples from healthy and LRTI infants using the R package *DESeq2* (Love et al., 2014) available via Bioconductor (Huber et al., 2015). We imported our unnormalized genus counts table compiled from PathoScope2 as a DESeqDataSet and ran the function DESeq, using a design model that included infants’ HIV exposure (from an HIV infected mother) as a covariate. For microbiome data, DESeq2 has been shown to return lower false discovery rates than other differential tests (Mcmurdie and Holmes, 2014), and performs particularly well for smaller experiments (Weiss et al., 2017).

To test whether the presence or absence of certain genera at the first sampled timepoint were associated with LRTI, we performed Fisher’s exact test to determine if healthy and LRTI infants are equally likely to have each genus in their NP microbiome. Because very low-abundance genera could be the result of spurious alignments or contamination, we explored both a high threshold (>1% relative abundance) and a low threshold (>0.1% relative abundance) for defining presence of a genus.

#### Beta diversity and clustering

We computed a Bray-Curtis dissimilarity matrix between samples using *vegan*’s vegdist function. When applied to relative abundance values, Bray-Curtis dissimilarity between two samples *i* and *j* is defined as 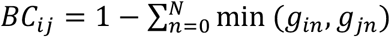 where *g*_*in*_ is the relative abundance of genus *n* in sample *i*. We performed hierarchical clustering of samples based on this dissimilarity matrix using R’s hclust function with the method set to “ward.D”. We defined clusters using R’s cuttree function, with the value for k selected by maximizing the Silhouette and Frey indexes as calculated by the package *NbClust* (Charrad et al., 2014). For each cluster, we performed Fisher’s exact tests to determine whether that cluster was enriched for LRTI samples generally, pre-symptomatic samples, active symptom samples, or HIV-exposed samples.

We used the metaMDS function from the R package *vegan* to perform non-metric multidimensional scaling (NMDS) ordination on our Bray-Cutris dissimilarity matrix, using as parameters k=3, try=50, and trymax=1000. Scaling our data onto just two dimensions using NMDS yielded a stress value greater than 0.2, indicating a poor fit; we instead scaled the data onto three dimensions (stress=0.13), and used the vegan’s envfit function to project the age and LRTI status of each sample into the NMDS ordination. 2-dimensional plots of our NMDS ordinated data can be found in **Figure 2**, and a 3-dimensional plot can be found in **Supplemental Figure 4**.

#### Differential analysis of maternal NP microbiomes

We used Spearman correlation coefficients to verify that the composition of infant NP microbiomes is related to their mother’s NP microbiome. We chose Spearman correlation, which utilizes rank order rather than continuous values, due to the compositional nature of bacterial abundance data. We calculated Spearman’s ρ for the relative abundance of each genus between mothers and their infants. We tested the significant of these correlations by comparing the distribution of ρ values to 1000 null distributions of the same metric, generated by randomly permuting the mother/infant labels.

We used DESeq2 to test for differential abundance of genera in the NP microbiomes of mothers of LRTI infants and mothers of control infants. For this analysis, we only included samples taken from mothers at the earliest pediatric visits, before their infants began exhibiting LRTI symptoms. We included the HIV status of the mothers as a covariate in DESeq2’s regression model. We report p-values after FDR correction via Benjamini-Hochberg procedure, and consider adjusted p-values below 0.1 to be significant.

## Data Availability

The raw and processed sequencing data from this study are available in the SRA repository, under accession number pending. Furthermore, all code, processed data, and the sample information metadata are available in the following GitHub repository: https://github.com/tfaits/Infant_Nasopharyngeal_Dysbiosis. Taxon counts tables are called "species.RDS", "genus.RDS", and "phylum.RDS". For strain/subspecies-level counts, "PathoScopeTable.txt" has the unfiltered/unprocessed outputs from PathoScope.

## Declarations

### Consent for publication

N/A

## Acknowledgments

Not applicable.

## Supplemental figures

**Supplemental Figure 1A).**
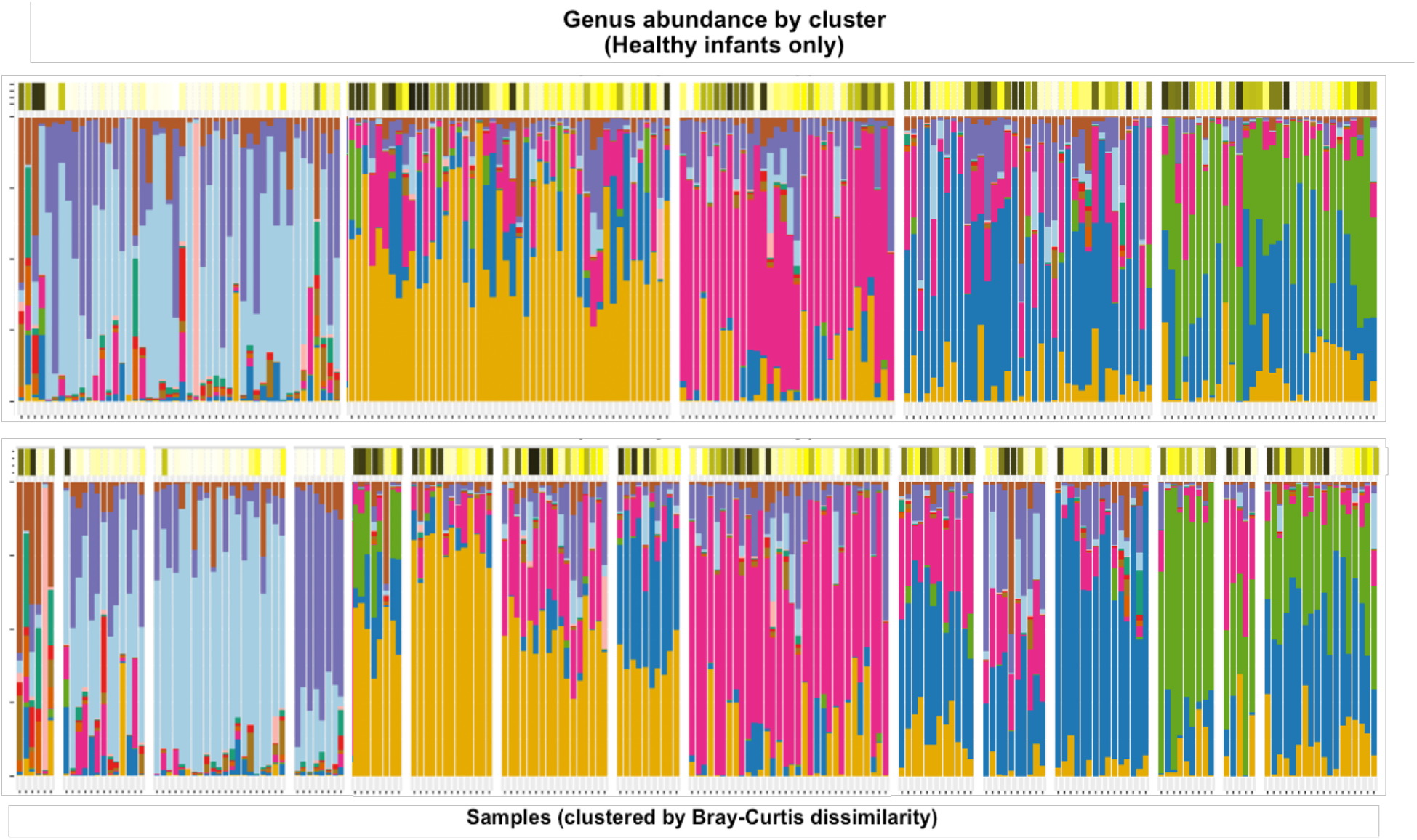
The taxonomic profiles of healthy infants’ NP samples (n=30), clustered by pairwise Bray-Curtis dissimilarity. Clusters were defined by performing hierarchical clustering on the beta diversity matrix and then cutting the resulting dendrogram into an optimal number of clusters according to the Silhouette index (5) and Frey index (15). The color bars above the stacked bar plots indicate the infants’ ages at the time of each sample.

**Supplemental Figure 1B).**
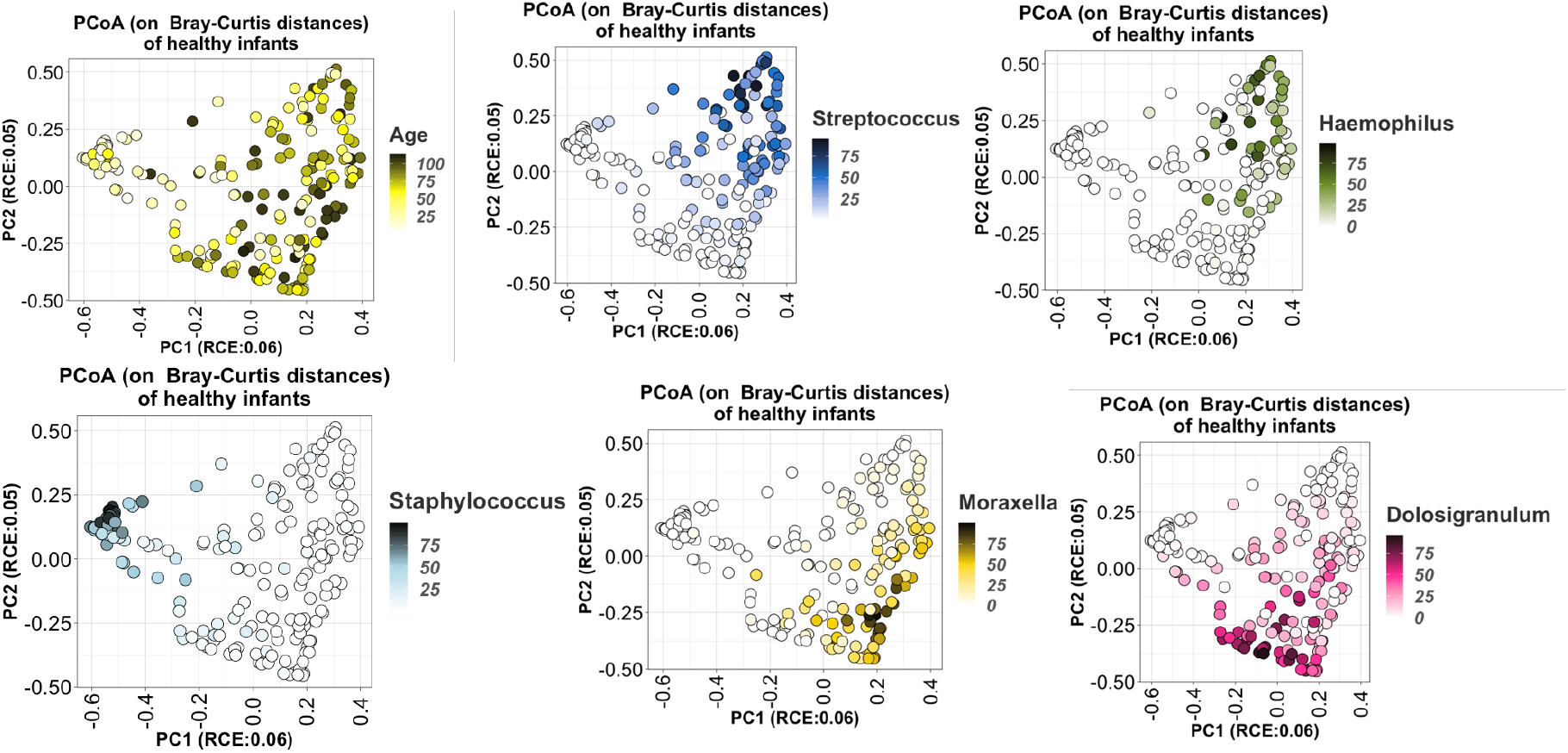
Principal Coordinate Analysis (PCoA) of the Bray-Curtis dissimilarity matrix between healthy infants’ samples, colored by age and relative abundance of the dominant genera.

**Supplemental Figure 2:**
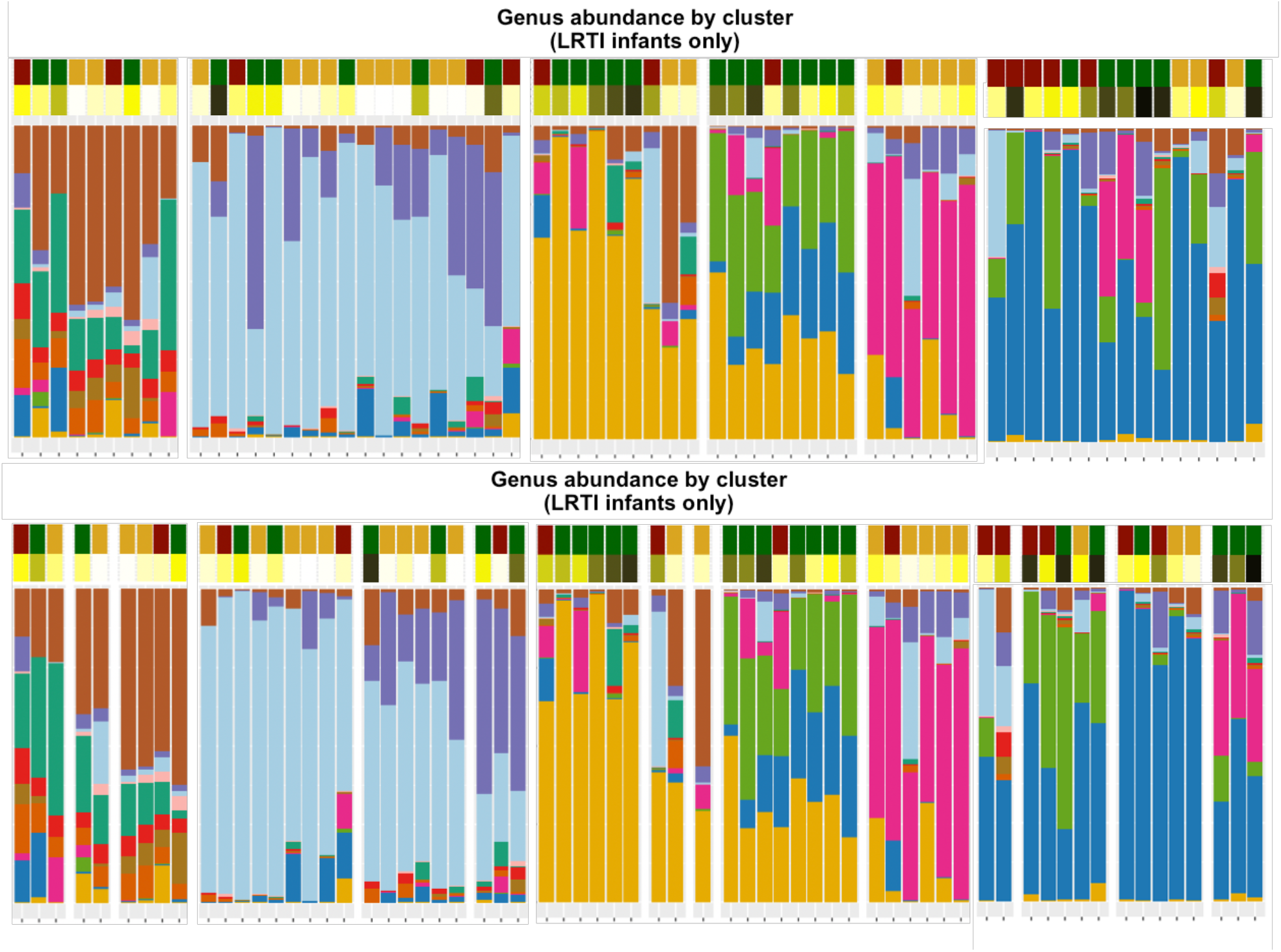
The taxonomic profiles of LRTI infants’ NP samples (n=10), clustered by pairwise Bray-Curtis dissimilarity. Clusters were defined by performing hierarchical clustering on the beta diversity matrix and then cutting the resulting dendrogram into an optimal number of clusters according to the Silhouette index (6) and Frey index (15). The color bars above the stacked bar plots indicate the infants’ ages and symptom status at the time of each sample.

**Supplemental Figure 3:**
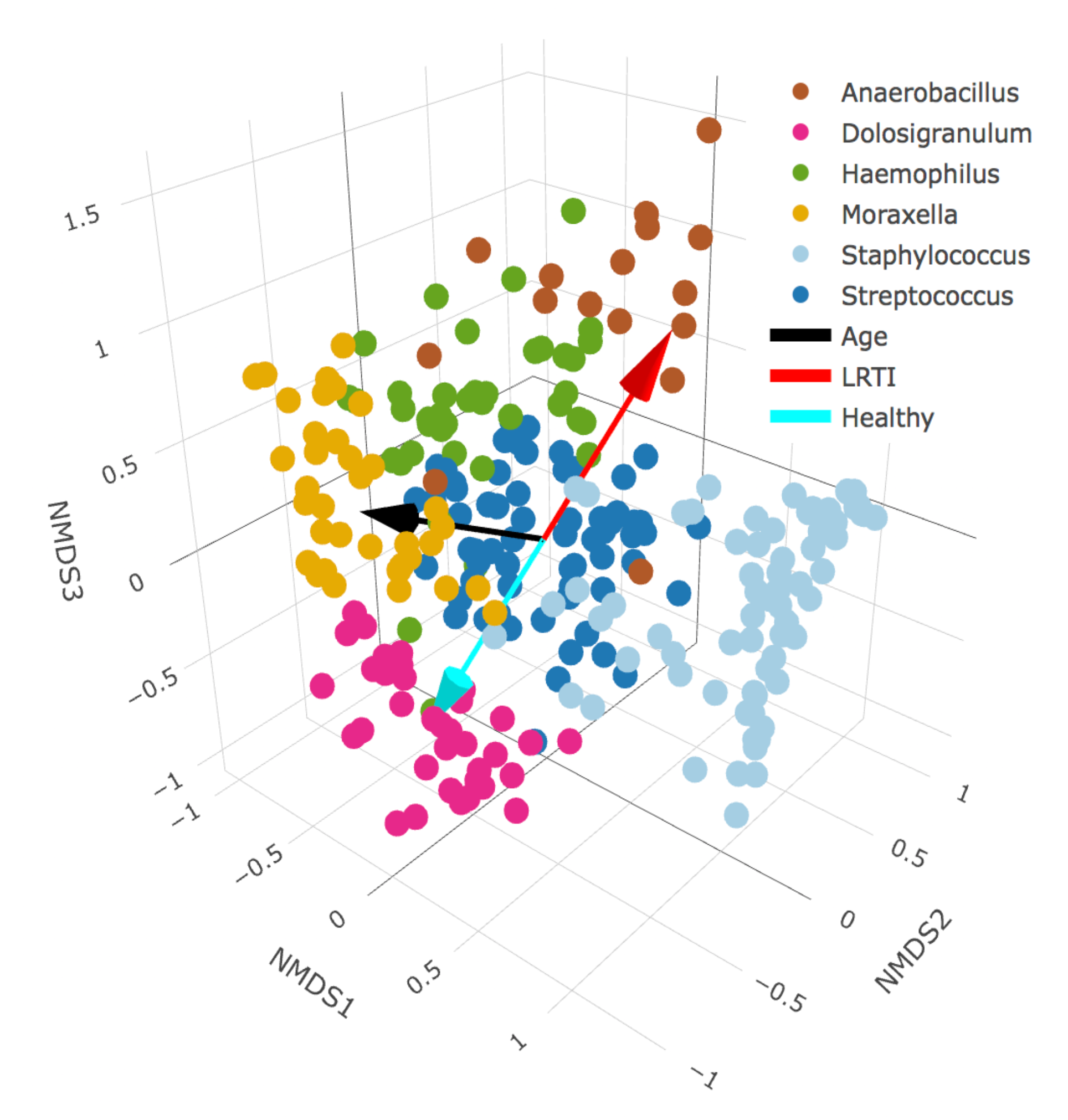
A 3-D scatterplot of infant nasopharyngeal samples projected into nonmetric multidimensional scaling (NMDS) ordination space. We applied 3-dimensional NMDS ordination to the Bray-Curtis dissimilarity matrix between all infants’ NP swabs, and projected vectors into that ordination space representing the best fit correlations for the age at sampling (the black vector) and LRTI status (the cyan vector represent control infants, the red vector represent LRTI infants). Samples are colored by their primary taxonomic profile cluster membership (see Figure 3 for details). Age is highly correlated with the x-axis, and samples on the young end of the age vector mostly belong to the *Staphylococcus*-dominated profile, whereas samples on the older end tend to belong more to the *Haemophilus* and *Moraxella*-dominated profiles. The *Dolosigranulum*-dominated profile is associated with the healthy end of the vector for LRTI status, while the *Anaerbacillus*-dominated profile is associated with disease.

**Supplemental Figure 4:**
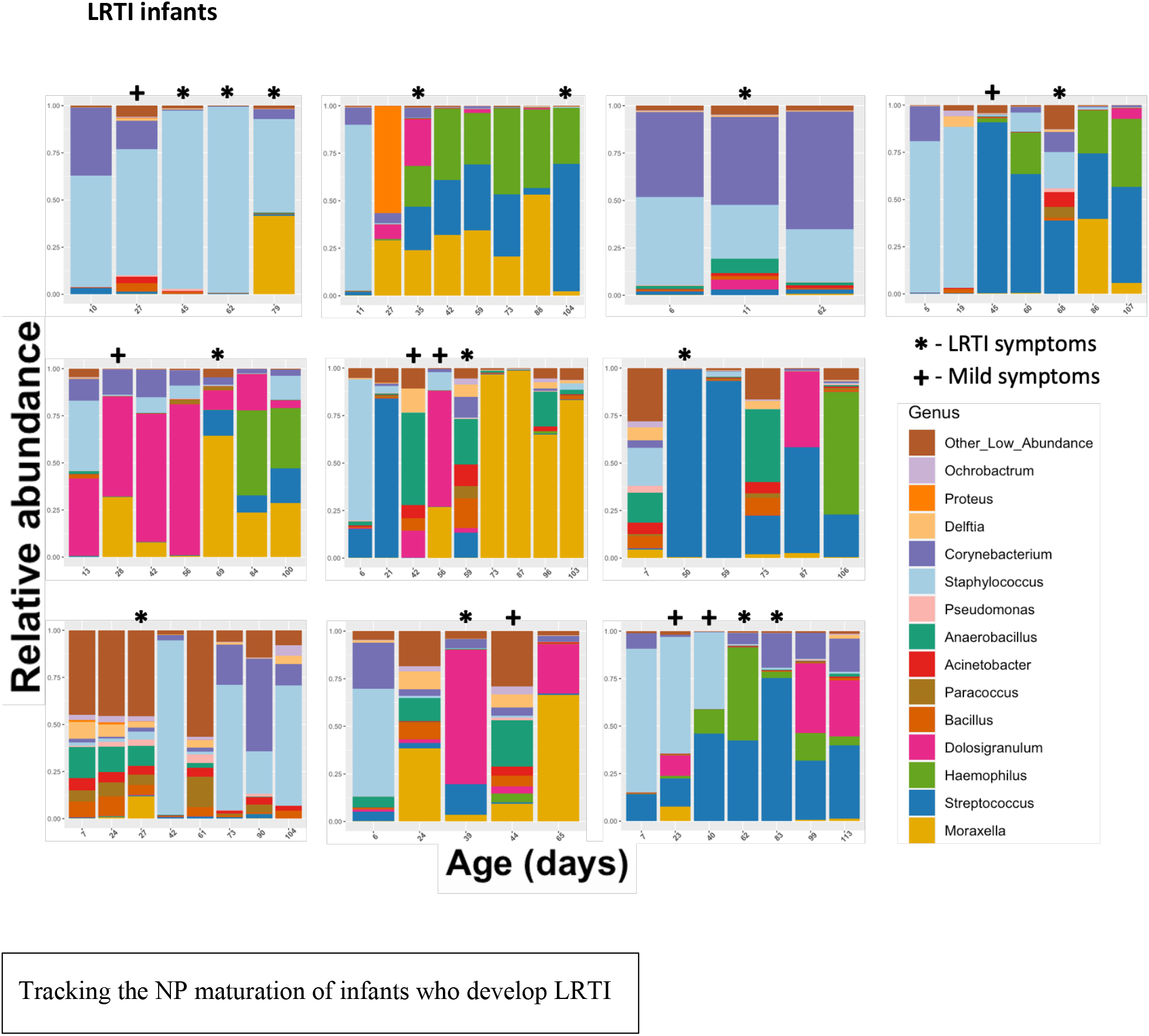
Each facet represents the relative abundance of bacterial genera in one infant’s nasopharynx over time. Samples are ordered chronologically from left to right, with labels on the x-axis displaying the infant’s age at the time of the NP swab. Time points where infants were experiencing severe respiratory symptoms are marked with a **\***. Time points where infants were experiencing mild symptoms which did not qualify as LRTI are marked with a **+**. Note that the order of these infants (reading rows left to right, top row first) is the same as the order in Table 2 (Clinical symptoms)

**Supplemental Figure 5:**
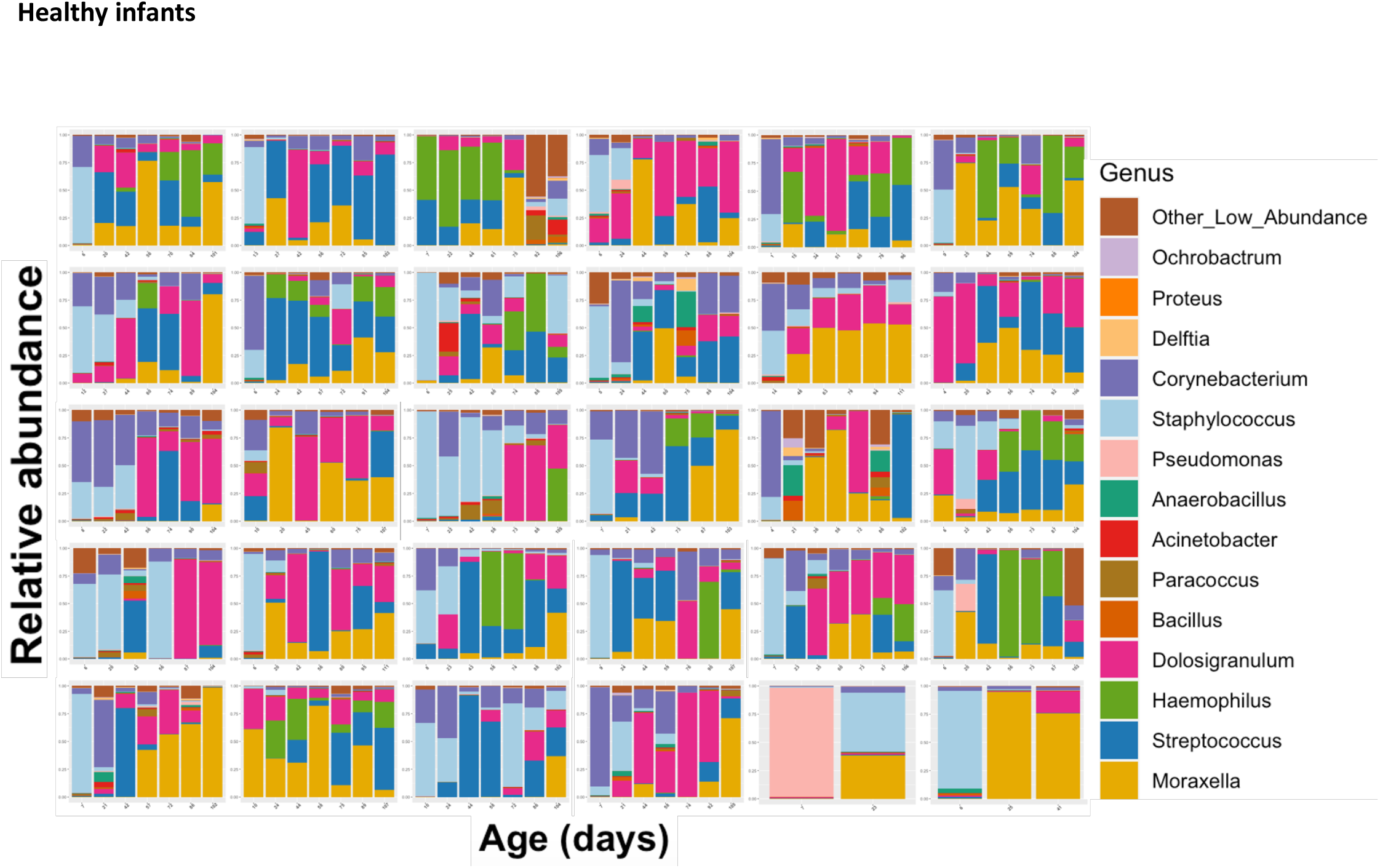
Tracking the NP maturation of healthy infants. Each facet represents the relative abundance of bacterial genera in one infant’s nasopharynx over time. Samples are ordered chronologically from left to right, with labels on the x-axis displaying the infant’s age at the time of the NP swab.

## References

Andrews S. 2010. FastQC. Babraham Bioinforma. https://doi.org/citeulike-article-id:11583827

Balsells E, Dagan R, Yildirim I, Gounder PP, Steens A, Muñoz-Almagro C, Mameli C, Kandasamy R, Givon Lavi N, Daprai L, van der Ende A, Trzciński K, Nzenze SA, Meiring S, Foster D, Bulkow LR, Rudolph K, Valero-Rello A, Ducker S, Vestrheim DF, von Gottberg A, Pelton SI, Zuccotti GV, Pollard AJ, Sanders EAM, Campbell H, Madhi SA, Nair H, Kyaw MH. 2018. The relative invasive disease potential of Streptococcus pneumoniae among children after PCV introduction: A systematic review and meta-analysis. J Infect 77:368–378. doi:10.1016/j.jinf.2018.06.004

Bates D, Mächler M, Bolker BM, Walker SC. 2015. Fitting linear mixed-effects models using lme4. J Stat Softw 67. doi:10.18637/jss.v067.i01

Benjamini Y, Hochberg Y. 1995. Controlling the False Discovery Rate: A Practical and Powerful Approach to Multiple Testing. J R Stat Soc Ser B 57:289–300. doi:10.1111/j.2517-6161.1995.tb02031.x

Biesbroek G, Bosch AATM, Wang X, Keijser BJF, Veenhoven RH, Sanders EAM, Bogaert D. 2014. The impact of breastfeeding on nasopharyngeal microbial communities in infants. Am J Respir Crit Care Med 190:298–308. doi:10.1164/rccm.201401-0073OC

Bokulich NA, Chung J, Battaglia T, Henderson N, Jay M, Li H, Lieber AD, Wu F, Perez-Perez GI, Chen Y, Schweizer W, Zheng X, Contreras M, Dominguez-Bello MG, Blaser MJ. 2016. Antibiotics, birth mode, and diet shape microbiome maturation during early life. Sci Transl Med 8. doi:10.1126/scitranslmed.aad7121

Bolger AM, Lohse M, Usadel B. 2014. Trimmomatic: A flexible trimmer for Illumina sequence data. Bioinformatics 30:2114–2120. doi:10.1093/bioinformatics/btu170

Bolyen E, Rideout JR, Dillon MR, Bokulich NA, Abnet CC, Al-Ghalith GA, Alexander H, Alm EJ, Arumugam M, Asnicar F, Bai Y, Bisanz JE, Bittinger K, Brejnrod A, Brislawn CJ, Brown CT, Callahan BJ, Caraballo-Rodríguez AM, Chase J, Cope EK, Da Silva R, Diener C, Dorrestein PC, Douglas GM, Durall DM, Duvallet C, Edwardson CF, Ernst M, Estaki M, Fouquier J, Gauglitz JM, Gibbons SM, Gibson DL, Gonzalez A, Gorlick K, Guo J, Hillmann B, Holmes S, Holste H, Huttenhower C, Huttley GA, Janssen S, Jarmusch AK, Jiang L, Kaehler BD, Kang K Bin, Keefe CR, Keim P, Kelley ST, Knights D, Koester I, Kosciolek T, Kreps J, Langille MGI, Lee J, Ley R, Liu YX, Loftfield E, Lozupone C, Maher M, Marotz C, Martin BD, McDonald D, McIver LJ, Melnik A V., Metcalf JL, Morgan SC, Morton JT, Naimey AT, Navas-Molina JA, Nothias LF, Orchanian SB, Pearson T, Peoples SL, Petras D, Preuss ML, Pruesse E, Rasmussen LB, Rivers A, Robeson MS, Rosenthal P, Segata N, Shaffer M, Shiffer A, Sinha R, Song SJ, Spear JR, Swafford AD, Thompson LR, Torres PJ, Trinh P, Tripathi A, Turnbaugh PJ, Ul-Hasan S, van der Hooft JJJ, Vargas F, Vázquez-Baeza Y, Vogtmann E, von Hippel M, Walters W, Wan Y, Wang M, Warren J, Weber KC, Williamson CHD, Willis AD, Xu ZZ, Zaneveld JR, Zhang Y, Zhu Q, Knight R, Caporaso JG. 2019. Reproducible, interactive, scalable and extensible microbiome data science using QIIME 2. Nat Biotechnol. doi:10.1038/s41587-019-0209-9

Bosch AATM, De Steenhuijsen Piters WAA, Van Houten MA, Chu MLJN, Biesbroek G, Kool J, Pernet P, De Groot PKCM, Eijkemans MJC, Keijser BJF, Sanders EAM, Bogaert D. 2017. Maturation of the infant respiratory microbiota, environmental drivers, and health consequences. Am J Respir Crit Care Med 196:1582–1590. doi:10.1164/rccm.201703-0554OC

Brugger SD, Bomar L, Lemon KP. 2016. Commensal–Pathogen Interactions along the Human Nasal Passages. PLOS Pathog 12:e1005633. doi:10.1371/journal.ppat.1005633

Cao B, Ho J, Retno Mahanani W, Louise Strong World Bank Group Emi Suzuki K, Andreev K, Bassarsky L, Gaigbe-Togbe V, Gerland P, Gu D, Hertog S, Li N, Spoorenberg T, Ueffing P, Wheldon M, Bay G, Cruz Castanheira H, Alkema L, Black R, Hopkins J, Guillot M, Hill K, Pedersen J, Jon Wakefield F, Liu L, Perin J, Villavicencio F, Yeung D, Ganesh Director V, Zhang Y, Hancioglu A, Avanesyan K, Bania S, Carter K, Carvajal L, Coskun Y, Delamónica E, Hanafy A, Hassfurter K, Jaques Y, Khan S, Kumapley R, Noeva R, Olivetti D, Quintana E, Ranck A, Requejo J, Unalan T, Young U. 2019. London School of Hygiene & Tropical Medicine Trevor Croft, The Demographic and Health Surveys (DHS) Program, ICF.

Charrad M, Ghazzali N, Boiteau V, Niknafs A. 2014. Nbclust: An R package for determining the relevant number of clusters in a data set. J Stat Softw 61:1–36. doi:10.18637/jss.v061.i06

de Steenhuijsen Piters WAA, Heinonen S, Hasrat R, Bunsow E, Smith B, Suarez-Arrabal M-C, Chaussabel D, Cohen DM, Sanders EAM, Ramilo O, Bogaert D, Mejias A. 2016. Nasopharyngeal Microbiota, Host Transcriptome, and Disease Severity in Children with Respiratory Syncytial Virus Infection. Am J Respir Crit Care Med 194:1104–1115. doi:10.1164/rccm.201602-0220OC

de Steenhuijsen Piters WAA, Jochems SP, Mitsi E, Rylance J, Pojar S, Nikolaou E, German EL, Holloway M, Carniel BF, Chu Mljn, Arp K, Sanders EAM, Ferreira DM, Bogaert D. 2019. Interaction between the nasal microbiota and S. pneumoniae in the context of live-attenuated influenza vaccine. Nat Commun 10:1–9. doi:10.1038/s41467-019-10814-9

de Steenhuijsen Piters WAA, Sanders EAM, Bogaert D. 2015. The role of the local microbial ecosystem in respiratory health and disease. Philos Trans R Soc B Biol Sci. doi:10.1098/rstb.2014.0294

Fischer Walker CL, Rudan I, Liu L, Nair H, Theodoratou E, Bhutta ZA, O’Brien KL, Campbell H, Black RE. 2013. Global burden of childhood pneumonia and diarrhoea. Lancet. doi:10.1016/S0140-6736(13)60222-6

Gill CJ, Mwananyanda L, MacLeod W, Kwenda G, Mwale M, Williams AL, Siazeele K, Yang Z, Mwansa J, Thea DM. 2016. Incidence of severe and nonsevere pertussis among HIV-exposed and-unexposed zambian infants through 14weeks of age: Results from the southern Africa mother infant pertussis study (samips), a longitudinal birth cohort study. Clin Infect Dis 63:S154–S164. doi:10.1093/cid/ciw526

Hasegawa K, Linnemann RW, Mansbach JM, Ajami NJ, Espinola JA, Petrosino JF, Piedra PA, Stevenson MD, Sullivan AF, Thompson AD, Camargo CA. 2017. Nasal Airway Microbiota Profile and Severe Bronchiolitis in Infants: A Case-control Study. Pediatr Infect Dis J 36:1044–1051. doi:10.1097/INF.0000000000001500

Hong C, Manimaran S, Shen Y, Perez-Rogers JF, Byrd AL, Castro-Nallar E, Crandall KA, Johnson WE. 2014. PathoScope 2.0: A complete computational framework for strain identification in environmental or clinical sequencing samples. Microbiome 2. doi:10.1186/2049-2618-2-33

Huber W, Carey VJ, Gentleman R, Anders S, Carlson M, Carvalho BS, Bravo HC, Davis S, Gatto L, Girke T, Gottardo R, Hahne F, Hansen KD, Irizarry RA, Lawrence M, Love MI, Macdonald J, Obenchain V, Oleś AK, Pagès H, Reyes A, Shannon P, Smyth GK, Tenenbaum D, Waldron L, Morgan M. 2015. Orchestrating high-throughput genomic analysis with Bioconductor HHS Public Access. Nat Methods 12:115–121. doi:10.1038/nmeth.3252

Ihekweazu FD, Versalovic J. 2018. Development of the Pediatric Gut Microbiome: Impact on Health and Disease. Am J Med Sci 356:413–423. doi:10.1016/j.amjms.2018.08.005

Kelly MS, Surette MG, Smieja M, Pernica JM, Rossi L, Luinstra K, Steenhoff AP, Feemster KA, Goldfarb DM, Arscott-Mills T, Boiditswe S, Rulaganyang I, Muthoga C, Gaofiwe L, Mazhani T, Rawls JF, Cunningham CK, Shah SS, Seed PC. 2017. The Nasopharyngeal Microbiota of Children with Respiratory Infections in Botswana. Pediatr Infect Dis J 36:e211–e218. doi:10.1097/INF.0000000000001607

Klindworth A, Pruesse E, Schweer T, Rg Peplies J, Quast C, Horn M, Glö Ckner FO. 2013. Evaluation of general 16S ribosomal RNA gene PCR primers for classical and next-generation sequencing-based diversity studies. Nucleic Acids Res 41. doi:10.1093/nar/gks808

Love MI, Huber W, Anders S. 2014. Moderated estimation of fold change and dispersion for RNA-seq data with DESeq2. Genome Biol 15:550. doi:10.1186/s13059-014-0550-8

Mansbach JM, Hasegawa K, Henke DM, Ajami NJ, Petrosino JF, Shaw CA, Piedra PA, Sullivan AF, Espinola JA, Camargo CA. 2016. Respiratory syncytial virus and rhinovirus severe bronchiolitis are associated with distinct nasopharyngeal microbiota. J Allergy Clin Immunol 137:1909-1913.e4. doi:10.1016/j.jaci.2016.01.036

Mcmurdie PJ, Holmes S. 2014. Waste Not, Want Not: Why Rarefying Microbiome Data Is Inadmissible. PLoS Comput Biol 10:1003531. doi:10.1371/journal.pcbi.1003531

Oksanen J, Blanchet FG, Friendly M, Kindt R, Legendre P, Mcglinn D, Minchin PR, O’hara RB, Simpson GL, Solymos P, Henry M, Stevens H, Szoecs E, Maintainer HW. 2019. Package “vegan” Title Community Ecology Package. Community Ecol Packag 2:1–297.

Revised WHO classification and treatment of childhood pneumonia at health facilities • EVIDENCE SUMMARIES •. n.d.

Stewart CJ, Ajami NJ, O’Brien JL, Hutchinson DS, Smith DP, Wong MC, Ross MC, Lloyd RE, Doddapaneni HV, Metcalf GA, Muzny D, Gibbs RA, Vatanen T, Huttenhower C, Xavier RJ, Rewers M, Hagopian W, Toppari J, Ziegler AG, She JX, Akolkar B, Lernmark A, Hyoty H, Vehik K, Krischer JP, Petrosino JF. 2018. Temporal development of the gut microbiome in early childhood from the TEDDY study. Nature 562:583–588. doi:10.1038/s41586-018-0617-x

Stewart CJ, Mansbach JM, Wong MC, Ajami NJ, Petrosino JF, Camargo CA, Hasegawa K. 2017. Associations of nasopharyngeal metabolome and microbiome with severity among infants with bronchiolitis: A multiomic analysis. Am J Respir Crit Care Med 196:882–891. doi:10.1164/rccm.201701-0071OC

Vavrek MJ. 2011. fossil: Palaeoecological and palaeogeographical analysis tools. Palaeontol Electron 14:16.

Weisberg JF and S. 2019. An R Companion to Applied Regression, Third. ed. Thousand Oaks (CA): SAGE Publications.

Weiss S, Xu ZZ, Peddada S, Amir A, Bittinger K, Gonzalez A, Lozupone C, Zaneveld JR, Vázquez-Baeza Y, Birmingham A, Hyde ER, Knight R. 2017. Normalization and microbial differential abundance strategies depend upon data characteristics. Microbiome 5:1–18. doi:10.1186/s40168-017-0237-y

Yildirim I, Hanage WP, Lipsitch M, Shea KM, Stevenson A, Finkelstein J, Huang SS, Lee GM, Kleinman K, Pelton SI. 2010. Serotype specific invasive capacity and persistent reduction in invasive pneumococcal disease. Vaccine 29:283–8. doi:10.1016/j.vaccine.2010.10.032

Yildirim I, Little BA, Finkelstein J, Lee G, Hanage WP, Shea K, Pelton SI. 2017. Surveillance of pneumococcal colonization and invasive pneumococcal disease reveals shift in prevalent carriage serotypes in Massachusetts’ children to relatively low invasiveness. Vaccine 35:4002–4009. doi:10.1016/j.vaccine.2017.05.077

